# The Eye-image Features of Patients with Coronary Heart Disease Assed: A prospective, observational study of traditional Chinese medicine combined with modern medicine

**DOI:** 10.1101/2023.07.26.23293223

**Authors:** Zhanqun Gao, Dirui Zhang, Ziqian Weng, Minghao Liu, Yubo Gao, Wei Hao, Chen Zhao, Ming Zeng, Xue Feng, Shengfang Wang, Boling Yi, Chunqi Xie, Yuhan Qin, Luping He, Yishuo Xu, Haibo Jia, Chao Fang, Zhi Zhang, Sining Hu, Bo Yu

**Author notes:** Zhanqun Gao, Dirui Zhang are co-first authors and have contributed equally to this work.

## Abstract

**BACKGROUND:** Coronary heart disease (CHD) significantly impacts human health. Traditional Chinese medicine (TCM) suggests a possible correlation between eye-image and CHD, but this relationship has not been fully explored in Western medicine.

**PURPOSE:** We aim to investigate the potential causal relationship between eye-image features and CHD, as examined by coronary angiography (CAG).

**METHODS:** The study selected patients hospitalized in the Department of Cardiology from November 15, 2021, to February 27, 2022. The selected patients were divided into two groups based on their CAG findings: the CHD group (at least one coronary lesion stenosis≥ 50%) and the control group (lesion stenosis<50%)

**RESURTS:** The final analysis included 342 patients out of a total of 426 participants, of these, 165 patients (48.2%) were diagnosed with CHD. The study found that certain characteristics in the left region 5(L5) and right region ( R5) were associated with CHD, including L5 pink dark speckle (OR: 4.143, 95%CI: 1.135-15.124, *P*=0.031), L5 vascular tortuosity (OR: 0.234, 95%CI: 0.077-0.71, *P*=0.010) R5 dark red blood vessels (known as Xue mai in TCM) (OR: 1.683, 95%CI: 1.035-2.738, *P*=0.036), and R5 yellowish mounds (OR: 2.083, 95%CI: 1.221-3.554, *P*= 0.007). Multivariate regression analyses showed that L5 vascular tortuosity had a negative correlation with CHD.

**CONCLUSION:** Our study revealed that four types of eye-image features, namely pink dark speckle, vascular tortuosity, dark red blood vessels, and yellowish mounds are associated with CHD. Among these features, vascular tortuosity showed a negative correlation with CHD, which could potentially aid in the diagnosis of the disease.

## Introduction

Coronary heart disease (CHD) is mainly caused by coronary atherosclerotic plaque or coronary artery spasm, leading to coronary artery stenosis, blood supply restriction, and myocardial ischemia and hypoxia[1]. Many studies have confirmed that CHD is the leading cause of cardiovascular death and chronic disability worldwide[2], becoming one of the health-threatening problems[3]. According to the World Health Organization, traditional Chinese medicine (TCM) is considered a branch of Western medicine that is used as an alternative or complementary treatment option worldwide[4], TCM is widely used in the daily lives of many Asians [5, 6], and has accumulated rich clinical experience[7]. Furthermore, TCM eye-image examination has the potential to be a valuable diagnostic tool for coronary heart disease (CHD) [8]. Previous studies have identified several eye-image features in the heart-eye region that are related to CHD according to TCM principles[9]. However, controversy exists due to differences in subjective or empirical factors among physicians and the lack of randomized, double-blind, placebo, and controlled clinical trials [10].

TCM believes that different parts of the sclera reflect the condition of different viscera. The color of speckles, mounds, and fogs in the eyes, as well as the color of blood vessels (called *Xue mai* in TCM) in the responding organ, can reflect the health of internal organs such as the lungs, kidneys, or heart [11]. According to the theory of TCM, 15 regions of each eye correspond to different 15 organs of the body. Particularly, the Left region 5 (L5) and Right region 5(R5) are believed to be related to the condition of the heart. These regions are located near the canthus of the eye, while the other side is closer to the pupil, as shown in Figure 1.

**Figure 1.**
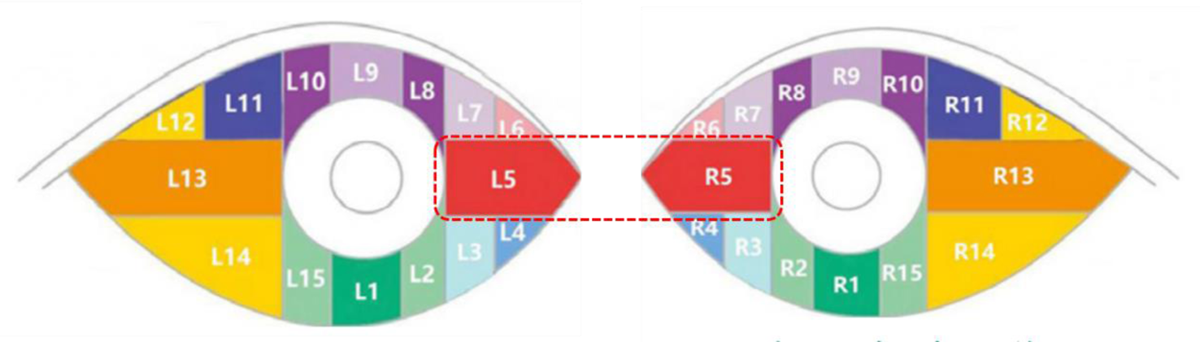
Schematic diagram of the relationship between the sclera and viscera L-left eye, R-right eye, 1, stomach; 2.15 spleen; 3, large intestine; 4, small intestine; 5, heart; 6, breast; 7, lung; 8, 10, kidney; 9, bladder; 11, reproductive system; 12, 14, liver; 13, gallbladder

Coronary angiography (CAG) is currently considered the most reliable method of diagnosing CHD [12]. However, it is an invasive and expensive procedure[13]. And traditional biomarker discoveries are not sufficient to provide a deeper understanding of the mechanisms implicated in CHD [14]. Therefore, in this study, a low-cost, simple, and novel method was used to evaluate CHD using eye-image features of integrated TCM and Western medicine.

## Methods

This paper was a single-center, retrospective, observational study consisting of 426 participants who were admitted to the Department of Cardiology at the Second Affiliated Hospital of Harbin Medical University (Harbin, China) between November 15, 2021, and February 27, 2022. The inclusion criteria for the study were patients aged 18-85 who were suspected of having CHD and were undergoing a CAG examination. Patients with a history of eye disease or surgery, such as infection, glaucoma, cataract, and so on, were excluded from the study. In addition, patients who wear colored contact lenses or artificial eyes are also excluded, Patients with PCI, CABG history as well as heart valve defects, and congenital heart defects are also excluded, as well as those whose eye-image pictures cannot be analyzed. Selected patients were divided into two groups according to angiographic stenosis[13]: the CHD group (at least one coronary artery stenosis of ≥50%), and the Control group(lesion stenosis <50%). A total of 342 eligible patients provided informed consent and participated in the research, undergoing both CAG and eye-imaging testing. The detailed study flowchart is illustrated in Figure 2.

**Figure 2.**
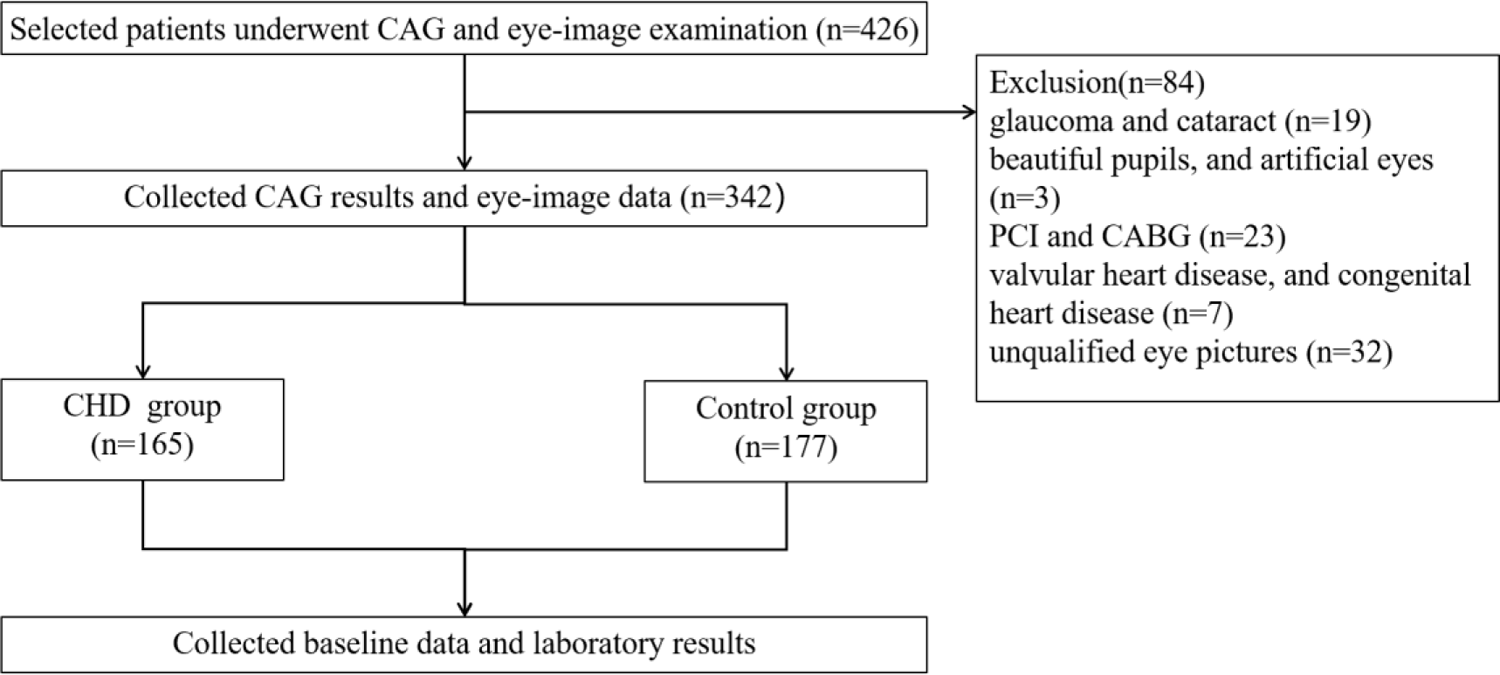
Study flowchart

### CAG examination

All selected patients were blinded to relevant studies via the femoral or radial artery by experienced interventional physicians. Using more than two orthogonal projection points to identify each lesion, stenosis grade was expressed as a percentage of coronary stenosis diameter and grouped according to coronary stenosis grade score.

### Eye-image system examination

The Capital Bio MyEyeD-10 system was utilized to examine eyes which combines the principles of both Chinese and Western medicine using advanced optical technology and artificial intelligence (AI). The system was additionally improved with the integration of the Askya imaging system, which has an impressive resolution of 28 million pixels.

The well-trained physician instructed the patient to place their eye on the small hole of the machine as shown in Figure 3A. Then the physician guided the patient to lift or pull down their eyelids with their finger to fully expose the sclera and rotate the direction of the eyeball. Once the light hit the sclera on the opposite side of the iris, the doctor quickly captured an image of the eye into the computer imaging system, this process was repeated for the other eye. Finally, eye-image were captured from five different angles (superior, inferior, left, right, and frontal) of each eye, as illustrated in Figure 3B, the analysis of the R5 and L5 regions was then collected.

**Figure 3.**
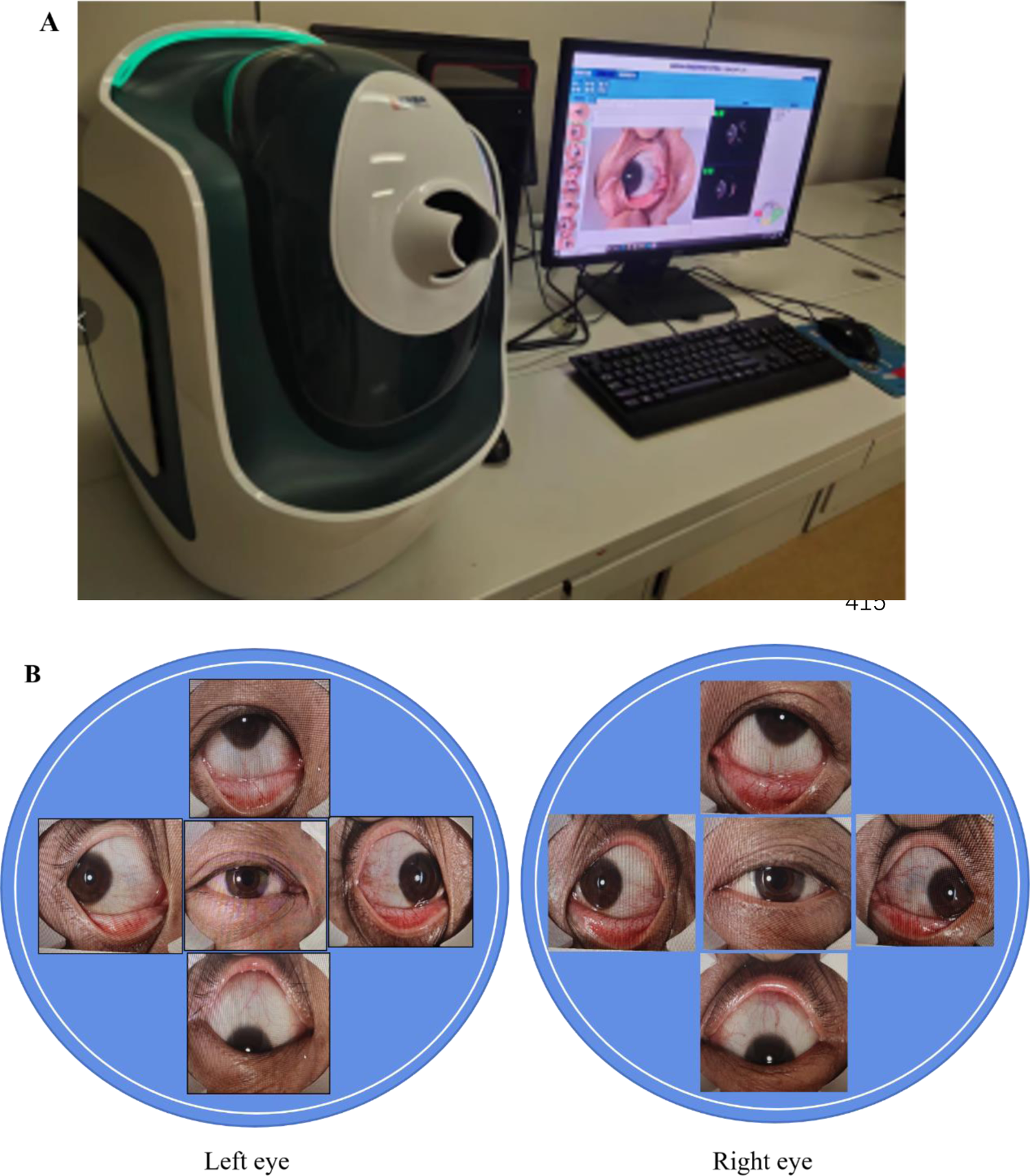
The eye-image system and taken pictures.

The eye-image system can automatically identify the color, shape, and size of the eye-image characteristics of patients with CHD. This is done by analyzing abnormal pathological phenomena in the heart-eye area, as well as determining whether the features protrude from the surface of the sclera. Based on these characteristics, the system can automatically name and judge the types of TCM syndromes that the patient may be experiencing. For example, speckles do not protrude from the surface of the sclera and are round, oval, or variously irregular in shape, and the pink dark speckle indicates blood stasis and blood deficiency. Mounds are round, oval, or irregular opaque bulges, with a diameter greater than 2mm, which is higher than the surface of the white eyes, and yellowish mounds are attributed to damp phlegm and blood stasis. Diseased blood vessels are no-red vessels of different lengths on the sclera, and dark red vessels are primarily caused by blood stasis. Vascular tortuosity is characterized by the repeated twisting and turning of blood vessels, regardless of color, and can display a continuous or intermittent state of contraction. This condition is primarily associated with Qi stagnation and blood stasis[15],. Pictures of eye-features as shown in Figure 4.

**Figure 4.**
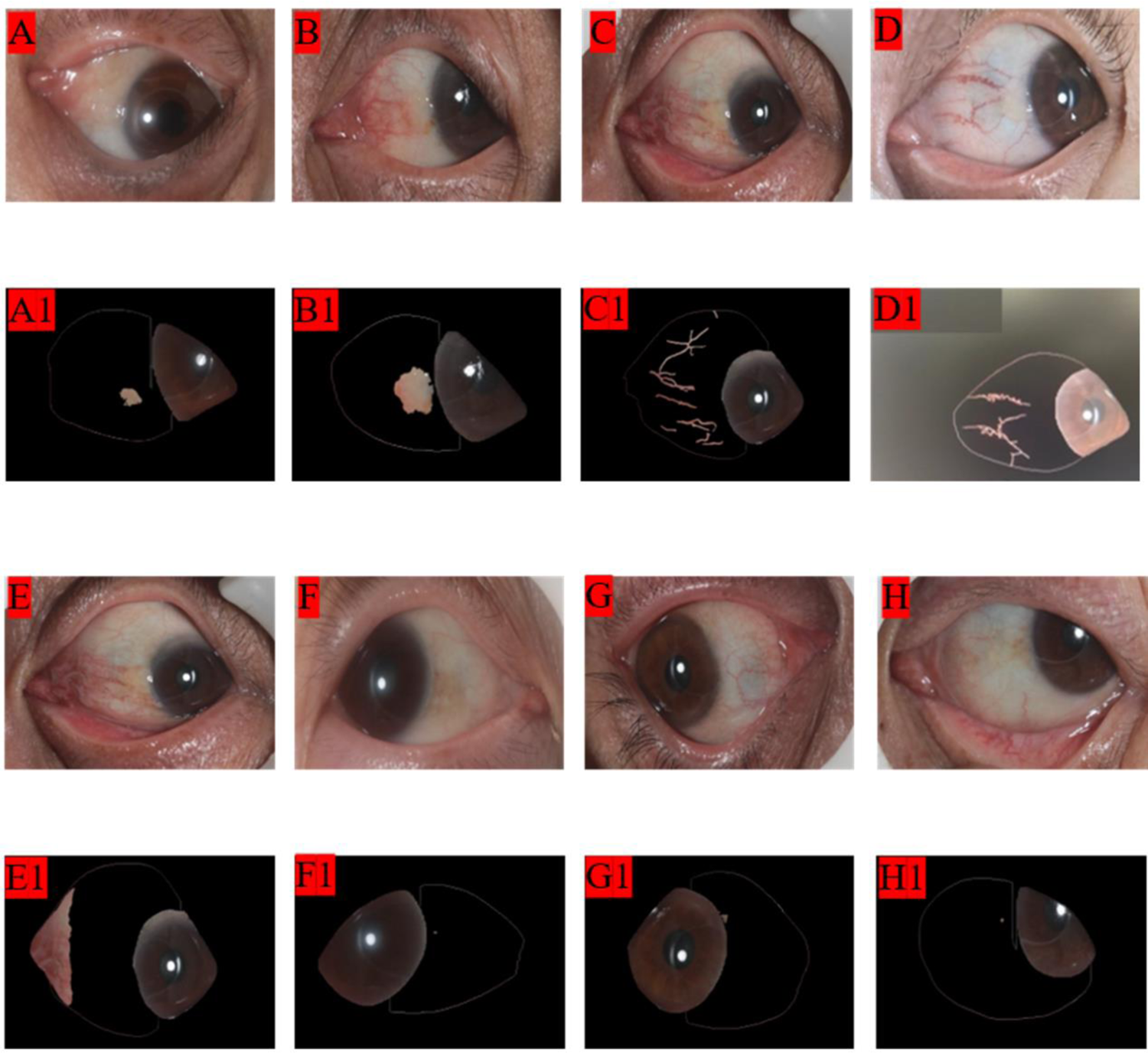
Eye-image forms and blood vessel A-H shows the taken eye-image pictures. A1-H1 shows analyzed eye-image forms in the eye-image system. A, A1, speckle, B, B1, mound, C, C1, blood vessel, D, D1, vascular tortuosity, E, E1, fog, F, F1, spot, G, G1, macula around black eye, H, H1, *Yue yun*

### Baseline data collection

To collect comprehensive medical history, information regarding diabetes, hypertension, stroke, and peripheral vascular disease (PVD) history. Additionally, it is important to obtain lifestyle habits including smoking history, including both past and present conditions. This should be accompanied by auxiliary examination results and laboratory values, such as ejection fraction (EF), glycated hemoglobin (HbAlc), creatinine (Cr), total cholesterol (T-Ch), total cholesterol (TG), HDL protein (HDL), low-density lipoprotein (LDL), and other relevant data.

### Statistical analysis

Statistical analysis was performed using SPSS V.25.0, continuous variables were presented as mean ± standard deviation, while categorical variables were presented as percentages. We also employed the chi-square test to compare categorical variables and the t-test to compare continuous variables. Additionally, we calculated exact 95% confidence intervals (CI) for all diagnostic performances. Univariate regression analysis and categorical variables were used to the comparison of the eye-image features in the CHD group and control group, and to analyze eye-image features using multivariate regression. In this study, *P*-values <0.05 was considered statistically significant.

## Results

From November 15, 2021, to February 27, 2022, a total of 426 patients underwent CAG and eye-image system examinations, 84 patients (19.7%) who did not meet the inclusion criteria were excluded, and 342 patients were included in the final analysis. Of all participants, 165 (48.2%) were diagnosed with CHD, of whom 116 (70.3%) were male, with a mean age of 58.7 ± 7.6 years. Additionally, 93 (56.4%) had hypertension, 66 (40.0%) smoked, and 38 (23.0%) had diabetes, and there were fewer cases of strokes and PVD (Table 1).

**Table 1.** Comparison of general data between the CHD group and the control group.

|  | Total(n=342) | CHD (n=165) | CG (n=177) | *P* value |
| --- | --- | --- | --- | --- |
| Male, n (%) | 192 (56.1) | 116 (70.3) | 76 (42.9) | < 0.001 |
| Age, year | 57.8±8.7 | 58.7±7.6 | 56.9±9.5 | < 0.001 |
| Diabetes, n (%) | 60 (17.5) | 38 (23.0) | 22 (12.4) | 0.010 |
| Hypertension, n (%) | 167 (48.8) | 93 (56.4) | 74 (41.8) | 0.007 |
| PVD, % | 11 (3.2) | 4 (2.4) | 7 (4.0) | 0.423 |
| Stroke, n (%) | 30 (8.8) | 19 (11.5) | 11 (6.2) | 0.083 |
| Smoking, n (%) | 98 (28.7) | 66 (40.0) | 32 (18.1) | < 0.001 |
| Serum glucose, mmol/ | 6.0±1.9 | 6.4±2.4 | 5.6±1.3 | 0.001 |
| HbA1c, % | 6.0±1.1 | 6.4±1.3 | 5.8±0.7 | 0.001 |
| Cr, umol/l | 70.9±35.8 | 72.9±30.7 | 69.1±39.8 | 0.322 |
| TG, mmol/L | 1.8±1.0 | 1.8±1.0 | 1.7±1.0 | 0.347 |
| HDL, mmol/L | 1.1±0.3 | 1.1±0.3 | 1.2±0.3 | < 0.001 |
| LDL, mmol/L | 2.9±0.9 | 3.0±1.0 | 2.8±0.8 | 0.283 |
| T-Ch, mmol/L | 4.4±1.0 | 4.4±1.1 | 4.5±0.93 | 0.672 |
| LVEF, % | 62.3±4.6 | 61.7±5.0 | 62.9±4.2 | 0.025 |

Values are mean SD, n (%), or mean (range). *p* values were determined by using an analysis of variance with a Bonferroni post hoc method for continuous variables and Fisher exact test for categorical variables.

In the study on eye-image features, we identified a total of 39 distinct features, including macula around the black eye, vascular tortuosity, and 6 colors of blood vessels, as well as various colors and shapes of speckles, mounds, spots, fogs, and Yue yuns. We further categorized these features into 8 colors of speckles, 7 colors of mounds, 6 colors of spots and fogs, and 4 colors of *Yue yuns* as shown in **Table 2**. An atypical colorless eye-image feature picture is shown **in Figure 4**. This study identifies 14 colors associated with CHD that appear in various forms and blood vessels. These colors include pink dark, dark pink, pink red, pink, dark red, red dark, red, yellow, dark yellow, pink yellow, pastel yellow, light yellow, and tan, as well as various light and grey. Our research indicates that colors associated with CHD are primarily compound colors, such as dark red and pink dark, as well as light yellow.

**Table 2.** Comparison of the eye-image features in the CHD group and control group.

| Eye-image features | L5 |  | *P-value* | R5 |  | *P-value* |
| --- | --- | --- | --- | --- | --- | --- |
|  | CHD (n=165) | CG (n=177) |  | CHD (n=165) | CG (n=177) |  |
| **Macula around black eye** | 42 (25.5) | 31 (17.5) | 0.073 | 38 (23.0) | 29 (16.4) | 0.122 |
| **Speckle** |  |  |  |  |  |  |
| Pink dark | 11 (6.7) | 3 (1.7) | 0.020 | 3 (1.8) | 2 (1.1) | 0.937 |
| Red dark | 5 (3.0) | 3 (1.7) | 0.647 | 8 (4.8) | 14 (7.9) | 0.249 |
| Yellow | 12 (7.3) | 15 (8.5) | 0.68 | 3 (1.8) | 3 (1.7) | 1.000 |
| Dark yellow | 9 (5.5) | 7 (4.0) | 0.512 | 9 (5.5) | 6 (3.4) | 0.351 |
| Light | 0(0) | 5 (2.8) | 0.085 | 6 (3.6) | 2 (1.1) | 0.24 |
| Grey | 3 (1.8) | 0(0) | 0.222 | 2 (1.2) | 0(0) | 0.448 |
| Red | 0(0) | 1 (0.6) | 0.334 | 2 (1.2) | 1 (0.6) | 0.951 |
| Pink-red | 1 (0.6) | 1 (0.6) | 1.000 | 0(0) | 0(0) | 1.000 |
| **Spot** |  |  |  |  |  |  |
| Pink dark | 6 (3.6) | 5 (2.8) | 0.671 | 2 (1.2) | 3 (1.7) | 1.000 |
| Red dark | 4 (2.4) | 2 (1.1) | 0.618 | 6 (3.6) | 4 (2.3) | 0.664 |
| Dark yellow | 8 (4.8) | 4 (2.3) | 0.194 | 1 (0.6) | 2 (1.1) | 1.000 |
| Grey | 2 (1.2) | 2 (1.1) | 1.000 | 3 (1.8) | 2 (1.1) | 0.937 |
| Light | 2 (1.9) | 2 (1.1) | 1.000 | 0(0) | 3 (1.7) | 0.272 |
| Red | 0(0) | 1 (0.6) | 1.000 | 0(0) | 0(0) | 1.000 |
| **Mound** |  |  |  |  |  |  |
| Yellowish | 45 (27.3) | 34 (19.2) | 0.077 | 45 (27.3) | 27 (15.3) | 0.006 |
| Light | 29 (17.6) | 31 (17.5) | 0.988 | 13 (7.9) | 11 (6.2) | 0.547 |
| Dark yellow | 14 (8.5) | 16 (9.0) | 0.856 | 6 (3.6) | 4 (2.3) | 0.45 |
| Grey | 1 (0.6) | 2 (1.1) | 1.000 | 5 (3.0) | 7 (4.0) | 0.642 |
| Pink yellow | 2 (1.2) | 5 (2.8) | 0.503 | 2 (1.2) | 4 (2.3) | 0.461 |
| Pink | 3 (1.8) | 2 (1.1) | 0.937 | 30 (18.2) | 35 (19.8) | 0.708 |
| Dark red | 1 (0.6) | 1 (0.6) | 1.000 | 1 (0.6) | 0(0) | 0.972 |
|  | CHD | CG |  | CHD | CG |  |
| **Blood vessel** |  |  |  |  |  |  |
| Pink dark | 74 (44.8) | 71 (40.1) | 0.376 | 54 (32.7) | 62 (35.0) | 0.653 |
| Dark red | 50 (30.3) | 39 (22.0) | 0.082 | 52 (31.5) | 38 (21.5) | 0.035 |
| Light | 0(0) | 2 (1.1) | 0.509 | 3 (1.8) | 7 (4.0) | 0.241 |
| Pink-red | 20 (12.1) | 19 (10.7) | 0.687 | 21 (12.7) | 19 (10.7) | 0.567 |
| Pink | 1 (0.6) | 4 (2.3) | 0.411 | 0(0) | 3 (1.7) | 0.272 |
| Red | 3 (1.8) | 10 (5.6) | 0.064 | 0(0) | 10 (5.6) | 0.005 |
| **Vascular tortuosity** | 4 (2.4) | 17 (9.6) | 0.006 | 6 (3.6) | 15 (8.5) | 0.063 |
| **Fog** |  |  |  |  |  |  |
| Pink dark | 104 (63.0) | 121 (68.4) | 0.299 | 116 (70.3) | 121 (68.4) | 0.697 |
| Red dark | 17 (10.3) | 21 (11.9) | 0.646 | 21 (12.7) | 12 (6.8) | 0.063 |
| Pink | 3 (1.8) | 2 (1.1) | 0.937 | 1 (0.6) | 0(0) | 0.972 |
| Yellow | 1 (0.6) | 0(0) | 0.972 | 1 (0.6) | 2 (1.1) | 1.000 |
| Dark yellow | 1 (0.6) | 1 (0.6) | 1.000 | 0(0) | 0(0) | 1.000 |
| Pink-red | 0(0) | 2 (1.1) | 0.509 | 0(0) | 1 (0.6) | 0.334 |
| **Yue yun** |  |  |  |  |  |  |
| Yellow | 1 (0.6) | 1 (0.6) | 1.000 | 0(0) | 0(0) | 1.000 |
| Dark pink | 0(0) | 0(0) | 1.000 | 0(0) | 1 (0.6) | 1.000 |
| Red dark | 0(0) | 0(0) | 1.000 | 1 (0.6) | 0(0) | 0.972 |
| Tawny | 0(0) | 0(0) | 1.000 | 1 (0.6) | 0(0) | 0.972 |

Note, Data presented as mean n (%), P -value < 0.05.

Interestingly, among single colors, red was found to be associated with the control group. We found that the four eye-image features related to CHD are all related to blood stasis, among them, the pink dark speckle, yellowish mound, and dark red blood vessel are positively correlated with CHD, while the tortuous blood vessel is negatively correlated with CHD.

After analyzing all the eye-image using the Capital Bio MyEyeD-10 system, we compared the differences in eye-image features between two groups of patients as shown in Table 2. Interestingly, we discovered that 5 eye-image features were significantly different between the two groups, as shown in Figure 5. There are L5 pink dark speckles (6.7% vs 1.7%, P=0.020), R5 dark red blood vessels (31.5% vs 21.5%, *P*=0.035), and R5 yellowish mound (27.3% vs 15.3%, *P*=0.006) and L5 vascular tortuosity (2.4% vs 9.6%, *P*=0.006) associated with CHD, and the R5 red blood vessel (0.0% vs5.6 %, *P*=0.005) is related with the control group.

**Figure 5.**
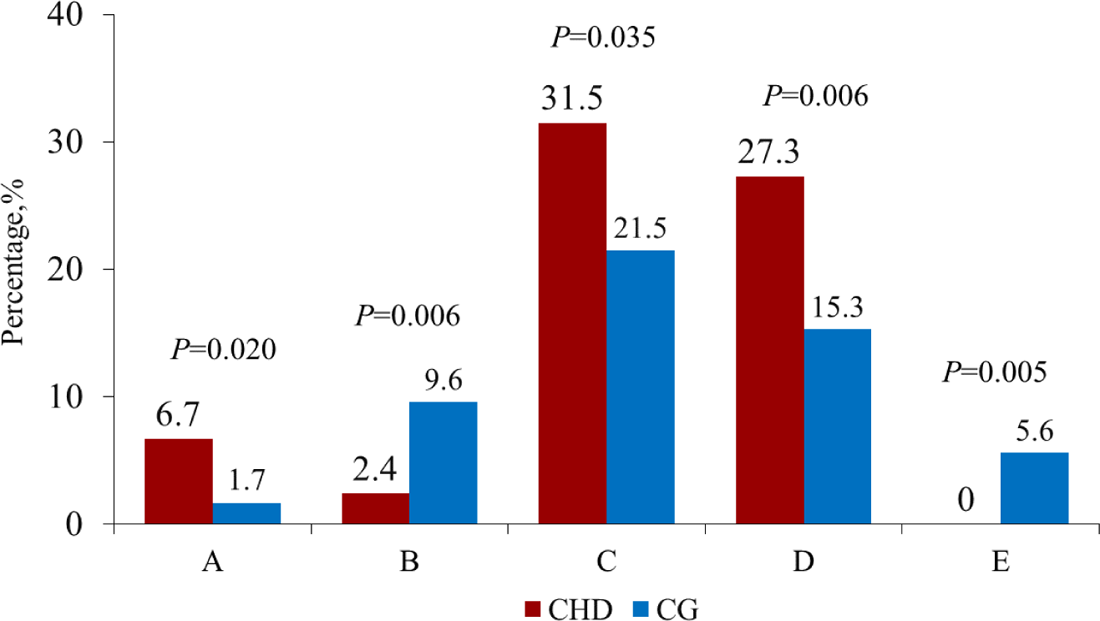
Meaningful eye-image features in the CHD group and CG(control group) A, pink dark speckle; B, L5 vascular tortuosity; C, R5 dark red blood vessel; D, R5 yellowish mound; E, R5 red blood vessel

The Univariate regression analysis results of eye-image features showed that there was a significant correlation between the 4 types of eye-image features and CHD, including L5 pink dark speckle (OR: 4.143, 95%CI: 1.135-15.124, *P*=0.031), L5 vascular tortuosity (OR: 0.234, 95%CI: 0.077-0.71, *P*=0.010), R5 dark red blood vessel (OR: 1.683, 95%CI: 1.035-2.738, *P*=0.036), and R5 yellowish mound (OR: 2.083, 95% CI:1.221 - 3.554, *P*=0.007), as shown in Table 3. In multivariate regression analysis was conducted to investigate the association between vascular tortuosity and CHD, while controlling for variables with statistical significance in a single factor and excluding the influence of collinearity. After adjusting for gender, age, smoking, and serum glucose, the results revealed that vascular tortuosity remained negatively associated with CHD. (Table 4)

**Table 3.** Comparison of the eye-image features in the CHD group and the control group.

| Eye-image features | L5 |  | *P value* | R5 |  | *P value* |
| --- | --- | --- | --- | --- | --- | --- |
|  | OR | 95% CI |  | OR | 95% CI |  |
| **Macula around the black eye** | 1.608 | 0.954-2.712 | 0.075 | 1.527 | 0.891-2.616 | 0.123 |
| **Speckle** |  |  |  |  |  |  |
| Pink dark | 4.143 | 1.135-15.124 | 0.031 | 1.620 | 0.267-9.821 | 0.600 |
| Red dark | 1.812 | 0.426-7.707 | 0.421 | 0.593 | 0.242-1.453 | 0.253 |
| Yellow | 0.847 | 0.384-1.868 | 0.681 | 1.074 | 0.214-5.398 | 0.931 |
| Dark yellow | 0.714 | 0.26-1.962 | 0.513 | 1.644 | 0.572-4.725 | 0.356 |
| Light | - | - | - | 3.302 | 0.657-16.596 | 0.147 |
| Grey | - | - | - | - | - | - |
| Red | - | - | - | 2.160 | 0.194-24.041 | 0.531 |
| Pink-red | 1.073 | 0.067-17.298 | 0.960 | - | - | - |
| **Spot** |  |  |  |  |  |  |
| Pink dark | 1.298 | 0.389-4.337 | 0.672 | 0.712 | 0.117-4.313 | 0.711 |
| Red dark | 2.174 | 0.393-12.029 | 0.374 | 1.632 | 0.452-5.890 | 0.454 |
| Dark yellow | 2.204 | 0.651-7.461 | 0.204 | 0.534 | 0.048-5.940 | 0.609 |
| Grey | 1.074 | 0.149-7.710 | 0.944 | 1.620 | 0.267-9.821 | 0.600 |
| Light | 1.074 | 0.149-7.720 | 0.944 | - | - | - |
| Red | - | - | - | - | - | - |
| **Mound** |  |  |  |  |  |  |
| Yellowish | 1.577 | 0.95-2.619 | 0.078 | 2.083 | 1.221-3.554 | 0.007 |
| Light | 1.004 | 0.575-1.754 | 0.988 | 1.291 | 0.561-2.967 | 0.548 |
| Dark yellow | 0.933 | 0.44-1.977 | 0.856 | 1.632 | 0.452-5.890 | 0.454 |
| Grey | 0.534 | 0.048-5.94 | 0.609 | 0.759 | 0.236-2.440 | 0.643 |
| Pink yellow | 0.422 | 0.081-2.206 | 0.307 | 0.531 | 0.096-2.936 | 0.468 |
| Pink | 1.620 | 0.267-9.821 | 0.600 | 0.902 | 0.525-1.550 | 0.708 |
| Dark red | 1.073 | 0.067-17.298 | 0.960 | - | - | - |
|  | OR | 95% CI |  | OR | 95% CI |  |
| **Blood vessel** |  |  |  |  |  |  |
| Pink dark | 1.214 | 0.79-1.865 | 0.376 | 0.902 | 0.576-1.413 | 0.653 |
| Dark red | 1.538 | 0.946-2.502 | 0.083 | 1.683 | 1.035-2.738 | 0.036 |
| Light | - | - | 0.999 | 0.450 | 0.114-1.769 | 0.253 |
| Pink-red | 1.147 | 0.589-2.235 | 0.687 | 1.213 | 0.627-2.347 | 0.567 |
| Pink | 0.264 | 0.029-2.384 | 0.235 | - | - | - |
| Red | 0.309 | 0.084-1.144 | 0.079 | - | - | - |
| **Vascular tortuosity** | 0.234 | 0.077-0.710 | 0.010 | 0.408 | 0.154-1.077 | 0.070 |
| **Fog** |  |  |  |  |  |  |
| Pink dark | 0.789 | 0.504-1.234 | 0.299 | 1.096 | 0.691-1.736 | 0.697 |
| Red dark | 0.853 | 0.433-1.681 | 0.646 | 2.005 | 0.953-4.218 | 0.067 |
| Pink | 1.620 | 0.267-9.821 | 0.600 | - | - | - |
| Yellow | - | - | - | 0.534 | 0.048-5.940 | 0.609 |
| Dark yellow | 1.073 | 0.067-17.298 | 0.960 | - | - | - |
| Pink red | - | - | - | - | - | - |
| **Yue yun** |  |  |  |  |  |  |
| Yellow | 1.073 | 0.067-17.298 | 0.960 | - | - | - |
| Dark pink | - | - | - | - | - | - |
| Red dark | - | - | - | - | - | - |
| Tawny | - | - | - | - | - | - |

Note, The L5 95% and R5 95% represent the 95% lower and upper limits of the OR, respectively. *P* -value < 0.05

**Table 4.** analysis of eye-image features using multivariate regressionsss.

| Variables | OR | 95%CI |  | *P value* |
| --- | --- | --- | --- | --- |
|  |  | lower limit | upper limit |  |
| L5- vascular tortuosity | 0.229 | 0.060 | 0.873 | 0.031 |
| Male | 2.966 | 1.727 | 5.095 | <0.001 |
| Age | 1.037 | 1.008 | 1.068 | 0.012 |
| Smoking | 0.256 | 0.256 | 0.829 | 0.010 |
| Serum glucose | 1.260 | 1.090 | 1.457 | 0.002 |

## Discussion

### Examination New Observation of CHD Patients

In this single-center, retrospective, and observational study, we identified significant differences in eye-image features between individuals with CHD and those without. Our analysis revealed four distinct eye-image features representing blood stasis associated with CHD, namely pink dark speckle, vascular tortuosity, dark red blood vessels, and yellowish mounds. Prior research indicates that a healthy person’s sclera is smooth and glossy, resembling the appearance of egg white, with no visible blood vessels. However, the onset of certain illnesses can alter this appearance, the sclera becomes abnormal, regardless of the severity of the disease or whether the body shows any symptoms[9, 11], and significant changes occur in the corresponding organ-eye region [4, 11] As the disease progresses, eye-image features form in connection with the ophthalmic veins or arteries on the surface or deep layer of the sclera[9]. In TCM, Yin-Yang and five-element theory mean the eye link to the heart[16] and other organs, also suggesting that the eyes are not an independent entity but are related to the human body’s internal environment [17]. Utilizing a non-invasive technique to extract eye-image characteristics from the heart-eye region can effectively diagnose the presence of CHD in a patient. This method is both cost-effective and straightforward and does not necessitate a specialized environment.

We demonstrated the validity of the TCM eye-image of CHD, which recognizes the human body as a unified whole[16] with interconnected organs and external manifestations of internal changes. For example, research has demonstrated that kidney disease can cause the thickening of the blood vessels in the kidney-eye area on the sclera, sometimes exceeding 0.24mm[11]. Our study conclusion shows a correlation between the eye-image features of the heart-eye area and CHD, which aligns with the holistic concept of TCM. Another important aspect is syndrome differentiation and treatment, as only accurate differentiation can lead to effective treatment. In TCM, CHD is diagnosed through patient *Zheng hou* (known as symptoms in Western medicine), which include cold and heat, deficiency and excess, phlegm dampness, and stasis [1, 18]. The disease *Zheng hou* of patients with CHD can be determined by analyzing the eye-image characteristics of the heart-eye area. This information can guide clinicians to choose appropriate treatment options using traditional Chinese or Western medicine.

### Eye-image features related to CHD

The four types of eye-image features related to CHD are all closely related to blood stasis, the source of blood stasis may be related to trauma, Qi and blood disorders, and pathogenic cold and heat[14]. Studies have shown that there is a certain correlation between blood stasis and the complexity of CHD, and the severity of blood stasis is an important factor for restenosis after percutaneous coronary intervention. It can be seen that blood stasis is closely related to patients with CHD[19]. Epidemiological studies in recent years have shown that stasis is the main syndrome of CHD, among 5284 patients with CHD, blood stasis accounted for 79.3%[7], In our study on eye-image features and their relation to CHD, yellowish mounds were the most commonly associated feature in Western medicine. These mounds are typically attributed to damp phlegm and blood stasis, as found in previous research[1, 18].

According to research [14], this condition is exacerbated by consuming large amounts of high-fat food and a lack of exercise, leading to the accumulation of lipids, increased blood lipid and blood cell adhesion, and the development of blood stasis[1]. It can be seen that the pathological process of blood stasis is consistent with the reason CHD is interlinked between Chinese and Western medicine[1, 14].

### *Zheng hou* and Eye-image features related to CHD

The research indicates that changes in the blood vessels of the eyes can evaluate the degree of microvascular and coronary lesions and also assess cardiovascular disease[20], to some extent, retinal vascular changes can also predict cardiovascular events[17]. The iris theory also supports that CHD is related to the eyes [21], by identifying the irregularity of the iris pigment between 2-3 points (roughly in the heart-eye area), it can be determined whether there is a lesion in the heart. Recently, applying a deep learning algorithm to analyze the eyes to develop CHD models to predict many cardiovascular risk factors and *Zheng hou* in TCM[22], with the help of AI and the combination of traditional Chinese[23] and Western medicine[24], the potential reason between TCM of AI[25] will become more and more clear in the future. Eye image features are inseparable from blood vessels, which can be seen on the sclera and connected to blood vessels deep in the sclera that cannot be seen with the naked eye [9]. Eye-image features may indicate panvascular disease or an extension of ocular atherosclerosis, but further research is needed to confirm these findings.

This study introduces a novel idea – that the vascular tortuosity on the heart-eye region of the sclera is negatively correlated with CHD. Previous research has established a negative correlation between retinal artery stenosis and CHD[26]. If we can confirm that the tortuous vascular structure is an artery and obtain precise measurements, we may be able to obtain more comprehensive and meaningful results. If it is confirmed that the tortuous blood vessels on the surface of the sclera are negatively correlated with coronary heart disease, it is more concise and convenient than detecting retinal arteries in the fundus.

Several studies have reported different levels of corneal changes in patients with CHD[26–28]. However, CHD is not only associated with the cornea but also with the sclera and retina of the eye. All three structures may contain similar markers that could help explain the link between CHD and the eyes. In this study, we found that the eye-image features of CHD patients were related to their left and right eyes, respectively. Chinese and Western medicine support research on the relationship between the left eye and the heart[9, 20], indicating that more research is needed.

### Limitations

The study has a small sample size and is limited to a single-center experiment. Future studies should include multicenter participation to avoid any deviations in the collection of eye-image features and data processing. Additionally, the study only collected static eye-images and did not consider dynamic attributes.

## Conclusion

Our study discovered distinct eye-image features in individuals with CHD, as compared to the control group. We propose a simple and feasible method that can aid in the screening of CHD during community physical examinations. If a patient exhibits typical eye image features, it may suggest a higher probability of CHD, and further examination is recommended.

## Authors contribution

Zhanqun Gao and Dirui Zhang: Conceptualization, Validation, Writing, Data analysis and statistical processing: Luping He, Yishuo Xu, Validation, Investigation, Writing: Ziqian Weng and Shengfang Wang, Data analysis: Boling Yi and Zhi Zhang, Data collection: Yubo Gao and Wei Hao, Data collection, Writing – original draft: Chunqi Xie, Yuhan Qin, Writing – review & editing: Ming Zeng, Xue Feng, Minghao Liu, and Chen Zhao: Draft revision, Haibo Jia and Chao Fang: Manuscript review and editing. Sining Hu and Bo Yu designed the whole experiment. All authors agree to accept full responsibility for all aspects of their work and to ensure completeness and accuracy.

## Abbreviation

AI: artificial intelligence

CAG: Coronary angiography

CABG: coronary artery bypass grafting

CHD: coronary heart disease

Cr: creatinine

EF: ejection fraction

HbAlC: glycosylated hemoglobin

HDL: high-density lipoprotein

LDL: low-density lipoprotein

TCM: traditional Chinese medicine

PCI: percutaneous coronary intervention

PVD: peripheral vascular disease

T-Ch: total cholesterol

TG: total triglyceride.

## Data Availability

We declare that there are no conflicts of interest in the publication and content of this manuscript.

